# Factorial Structure of the Transactional eHealth Literacy Among Vietnamese Youth: An Instrument Validation Study

**DOI:** 10.1101/2021.08.06.21261688

**Authors:** Giang Thu Vu, Thuc Minh Thi Vu, Robin van Kessel, Brian Li Han Wong, Tham Thi Nguyen, Thao Phuong Thi Nguyen, Son Hoang Nguyen, Ha Ngoc Do, Bach Xuan Tran, Carl A. Latkin, Cyrus Ho, Roger Ho

## Abstract

The progression into the Digital Age has brought an array of novel skill requirements. Unlike traditional literacy, there are currently few measures that can reliably measure eHealth literacy. The Transactional Model of eHealth Literacy and subsequent Transactional eHealth Literacy Instrument may provide a feasible option for measuring eHealth literacy. However, this instrument has yet to be validated, which is the aim of this study. We conducted an online cross-sectional study among 236 Vietnamese young people. Using exploratory factor analysis, we ensured that a model consisting of four factors had the best fit (RMSEA = 0.116; CFI = 0.907) and the highest internal consistency (Cronbach’s *α* = 0.96). A confirmatory factor analysis tested measurement invariance at four levels: configural, metric, scalar, and strict invariance. Only metric invariance was partially invariant, while the rest tested fully invariant. Even with partial metric invariance, there is reason to assume that functional, communicative, critical, and translational eHealth literacy (the four levels according to the transactional model) are consistently measured when deploying the Transactional eHealth Literacy Instrument across groups. In other words, this study suggests the instrument can be used for comparisons across groups and has the potential to generate high-quality data usable for informing change agents as to whether a particular population is proficient enough to adopt novel eHealth innovations.

## Introduction

eHealth literacy is a skill set that is becoming increasingly relevant as the Digital Age progresses (Sonnier, 2017). When healthcare and public health services slowly moved to the digital sphere (Odone et al., 2019), the skills required to search for, identify, and access these services started changing incrementally. However, much like most of the world, these services transitioned to digital environments at the beginning of the COVID-19 pandemic (Almeida et al., 2020). While innovations are typically taken up gradually by a population (Rogers, 1995), society adapted acutely to the pandemic - resulting in digital and technological advances neither equally nor equitably permeating all layers of society across the globe and potentially widening existing inequalities (Honeyman et al., 2020).

eHealth literacy is a specific branch in the field of digital literacy. To this day, it remains unclear what digital literacy is, how it can be assessed, and the benefits and risks from (not) being digitally literate (Chetty et al., 2018). UNESCO frames digital literacy as a set of foundational skills needed to work in the digital world and is considered a catalyst for individuals to achieve health and social outcomes (UNESCO, 2011). Current training programs tend to focus heavily on technical skills required to use digital technologies, while cognitive and ethical considerations are disregarded entirely (Chetty et al., 2018). It takes approximately five years for the average individual to become proficient in using digital technologies (Goldstuck, 2010). However, this five-year gap is not equal across all societal domains. For example, the medical sector has been slower to adapt to the digital world (Bartlett et al., 2021).

Recent work by Paige and colleagues created a tool with which to measure eHealth literacy (Paige et al., 2019). The Transactional eHealth Literacy Instrument (TeHLI) is a multi-dimensional instrument to measure increasing levels of eHealth literacy, based on the Transactional Model of eHealth Literacy (TMeHL) (Paige et al., 2018): functional (the ability to successfully read and write about health using technological devices), communicative (the ability to control, adapt, and collaborate communication about health with others in online social environments), critical (the ability to evaluate the relevance, trustworthiness, and risks of sharing and receiving health-related information on the Internet), and translational eHealth literacy (the ability to apply health-related information from the Internet in different contexts). However, research is needed to test the TeHLI with younger adults (Paige et al., 2019). This article aims to validate the TeHLI to see which components of the tool (how many components and which components included) would be the best fit statistically and whether the tool can be applied to groups of different characteristics (i.e. whether the different characteristics in terms of gender, education level, age group and other sociodemographic factors impact the effectiveness of the tool in any way).

## Materials and Methods

### Participants and study procedures

An online cross-sectional study was conducted among Vietnamese young people from April to June 2020 in Vietnam. The eligibility criteria for participating in this survey were: (1) ages between 16 and 35 years; (2) currently living in Vietnam; and (3) agreed to join this study by providing online informed consent. We recruited participants from all provinces of Vietnam using the snowball sampling technique. First, we developed a core group of participants (including leaders of the Youth Union in different public institutions, companies, and organisations) and invited them to complete the survey about the quality of life among youths. After completing the survey, these participants were requested to invite their peers in their respective networks to complete the online survey. At the end of the data collection period, 236 youths aged 16-35 living in Vietnam agreed to participate and completed the survey.

### Measurement and instrument

In this study, we built an online survey on the Survey Monkey platform. This approach is low cost, consumes little time, is user-friendly for youth, and is highly accessible to reach the samples nationwide. Each participant spent 5-10 minutes completing the survey. A structured questionnaire was used to collect information, consisting of two components: (1) sociodemographic characteristics and health status, and (2) the TeHLI. The survey was first piloted on five youths to ensure the cross-cultural validity of translated instruments in Vietnamese. After that, the revised questionnaire was uploaded into the online survey portal. The data collection began once the online survey system was tested to assure accurate question contents and no technical issues.

#### Sociodemographic and health status characteristics

Participants reported their sociodemographic information, including age, sex (male/female), educational attainment (below high school and high school/college/tertiary and higher), marital status (single/other), and living areas (urban/town/rural or mountain area) and acute symptoms in the last four weeks, any chronic conditions that they were diagnosed in the past.

##### Transactional eHealth Literacy Instrument

The TeHLI consisted of 18 items to reflect four aspects of the TMeHL such as functional (4 items), communicative (5 items), critical (5 items), translational (4 items). On a 5-point scale from 1 (strongly disagree) to 5 (strongly agree) was used for each item (Paige et al., 2019). The Cronbach’s *α* of four domains of TeHLI were 0.91; 0.92; 0.88; 0.92, respectively.

### Statistical analysis

Statistical analysis was performed using STATA version 16 and R. Standard descriptive statistical analysis was conducted with mean and standard deviation (SD) for quantitative variables and frequency and percentage for qualitative variables. The value of Skewness and Kurtosis coefficients were reported. Floor and ceiling effects were identified if the percentage of participants answering the lowest or highest response option was above 15% (McHorney & Tarlov, 1995). A p-value (p) <0.05 was considered statistically significant. The Exploratory Factor Analysis (EFA) using principal component analysis (PCA) was performed to indicate the items belonging to the four domains of the TMeHL: Model 1 (18 items divided into one factor); Model 2 (18 items into two factors); Model 3 (18 items into three factors); Model 3 (18 items into four factors). To determine the factor structure, we conducted a Confirmatory Factor Analysis (CFA) to test the original model (4 factors), then the new models of TeHLI. Multiple model fit indicators with respective cutoffs assessed the model fit of observed data (with Satorra-Bentler correction for non-normality data) (Hooper et al., 2008), including:

- Relative Chi-square (χ2 /df): a value ≤3.0 for good fit;
- Root Mean Square Error of Approximation (RMSEA): a value of ≤ 0.08 for good fit;
- Comparative Fit Index (CFI): a value of ≥ 0.9 for acceptable fit;
- Standardised Root Mean Square Residual (SRMR): a value of ≤ 0.08 for good fit

#### Measurement invariance

To determine the measurement invariance of the TeHLI, we performed a CFA by four increasingly constrained nested models. First, we evaluated *configural* invariance to assess what model is least stringent. This step aimed to test whether the constructs have the same pattern of free and fixed loadings across groups. Next, when configural invariance was supported, we evaluated *metric* invariance (also called pattern or weak invariance) or equivalence of the item loadings on the factors. Metric invariance was established by constraining factor loadings to be equivalent across groups. Next, we investigated *scalar* invariance by constraining equal intercepts and factor loadings across groups. This step assures that participants in different groups, on average, rate items similarly. Finally, we analysed *strict* invariance, which is supported when equal error variances are constrained in addition to equal intercepts and factor loadings. Testing residual error establishes that the same amount of error, or variance not accounted by the factor, is consistent for each item across groups. Measurement invariance is tested by some goodness of fit indexes, such as ΔCFI, ΔRMSEA, Δχ2. When we used Δχ^2^ as a sole measure of exact fit, the results showed that it was associated with lower levels of scalar invariance. By contrast, using the ΔCFI (with or without other criteria like ΔRMSEA) was associated with higher levels metrics, scalar, and strict model. Therefore, in this study, change in the CFI (ΔCFI) was used as the primary criterion for comparing models, and ΔCFI < .01 between successively more restricted models provides evidence for measurement invariance (Hu & Bentler, 1999; Putnick & Bornstein, 2016; Villarreal-Zegarra et al., 2019).

## Results

Table 1 showed the sociodemographic and health characteristics of the 236 participants in this study. The mean age of respondents was 21.3 ± 4.8, with the most common age group about above 24 years old (78.8%). The majority of those were female (71.2%). There was about 56.8% of participants had the education tertiary and upper. Regarding health status, about 44.5% and 16.5% of observations suffered from acute symptoms within the last four weeks and chronic conditions within the previous three months, respectively.

**Table 1.**
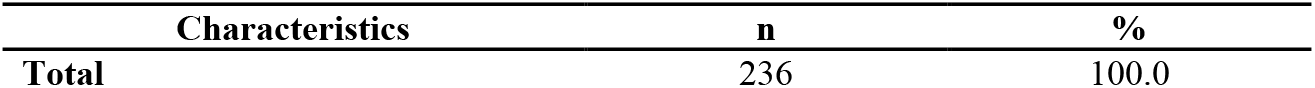
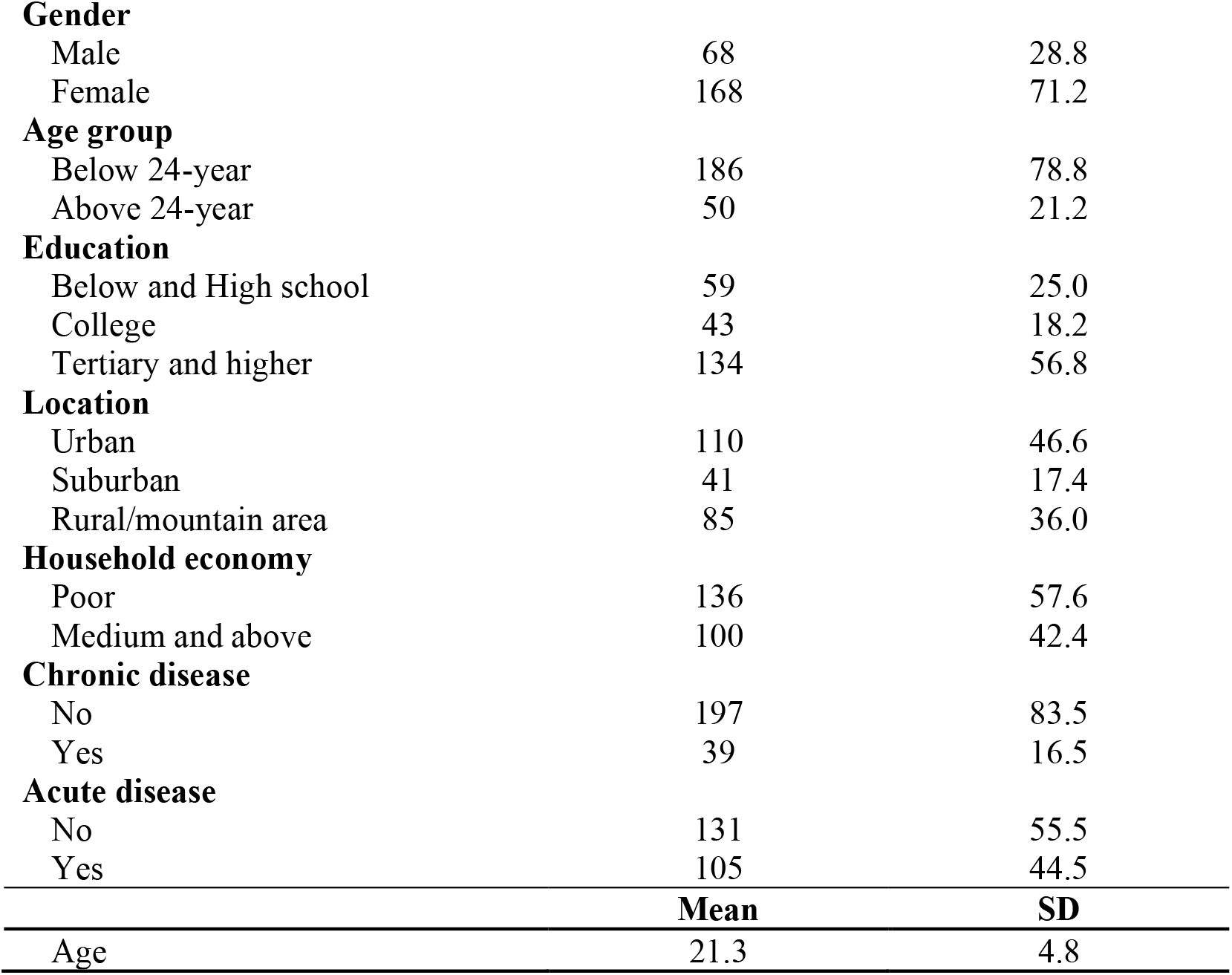
Demographic characteristics of participants.

Table 2 showed the results of the descriptive analysis for 18 items of the TeHLI. All of the 18 items had a range of scores from 1 to 5. Skewness and Kurtosis coefficients were reported with ranging from –1.0 to -0.2 and 2.8 to 4.3, respectively. Table 3 also presented the reliability of the modified TeHLI. Correlation coefficients with other items in respective factors ranged between 0.7 and 0.8. Internal consistency of factor 1 “Functional”, factor 2 “Communicative”, factor 3 “Critical”, and factor 4 “Translational” were good at 0.82, 0.92, 0.92, and 0.90, respectively. Table 3 further showed the results of Confirmatory Factor Analysis on four different models. The model with 1 factor of the TeHLI showed an RMSEA and CFI score of 0.157 and 0.806. In contrast, the model with four factors showed an RMSEA and CFI score of 0.116 and 0.907, respectively. As such, this model was used to conduct measurement invariance.

**Table 2.**
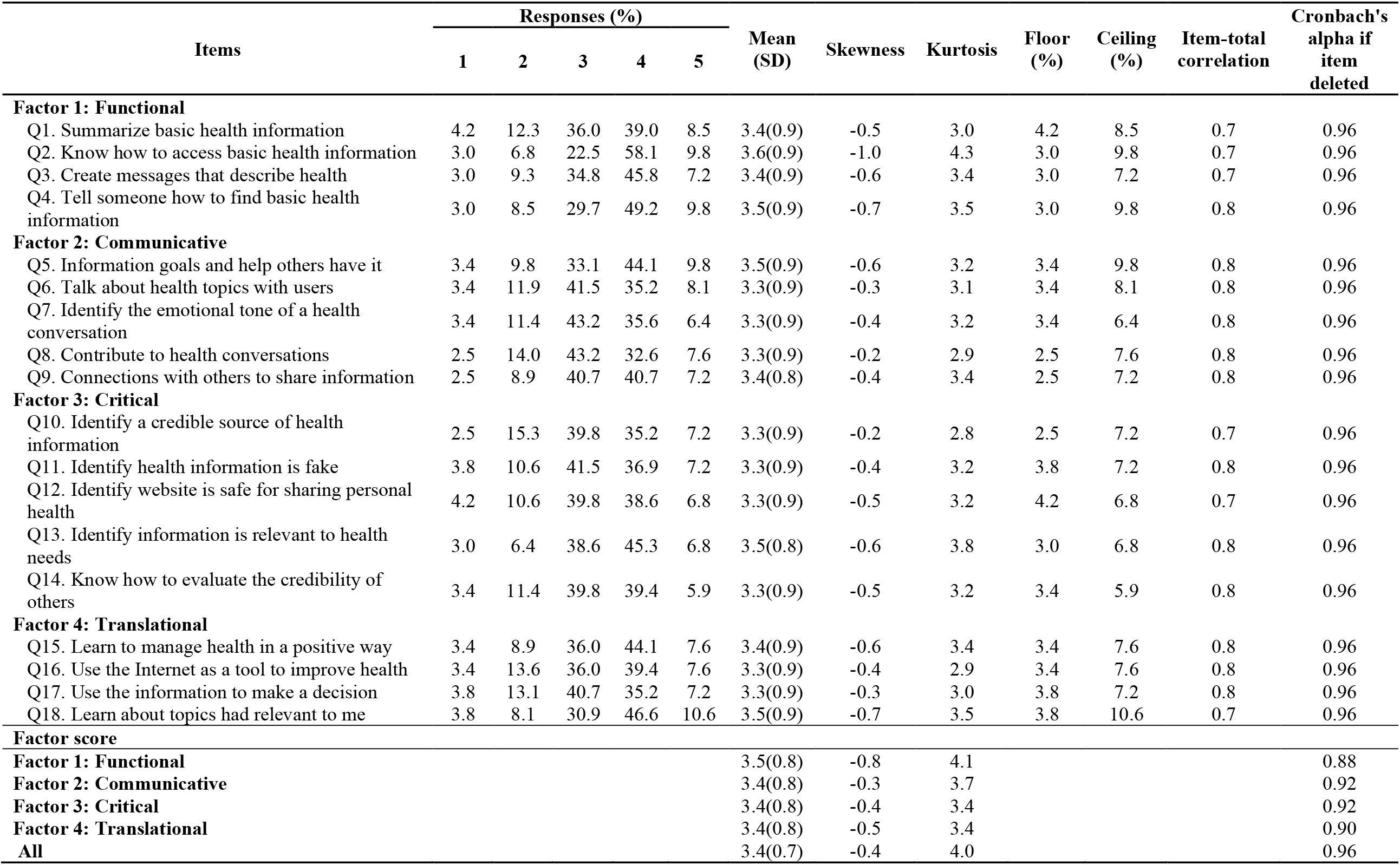
Basic descriptions and reliability of Transactional eHealth Literacy.

**Table 3.** Confirmatory Factor Analysis Models and Fit Indices Evaluating Factor Structure of the Transactional eHealth Literacy scale.

|  | **Model 1** | **Model 2** | **Model 3** | **Model 4** |
| --- | --- | --- | --- | --- |
| Q1. Summarise basic health information | ehealth literacy | Functional/Communicative | Functional | Functional |
| Q2. Know how to access basic health information | ehealth literacy | Functional/Communicative | Functional | Functional |
| Q3. Create messages that describe health | ehealth literacy | Functional/Communicative | Functional | Functional |
| Q4. Tell someone how to find basic health information | ehealth literacy | Functional/Communicative | Functional | Functional |
| Q5. Information goals and help others have it | ehealth literacy | Functional/Communicative | Communicative | Communicative |
| Q6. Talk about health topics with users | ehealth literacy | Functional/Communicative | Communicative | Communicative |
| Q7. Identify the emotional tone of a health-related conversation | ehealth literacy | Functional/Communicative | Communicative | Communicative |
| Q8. Contribute to health conversations | ehealth literacy | Functional/Communicative | Communicative | Communicative |
| Q9. Connections with others to share information | ehealth literacy | Functional/Communicative | Communicative | Communicative |
| Q10. Identify a credible source of health information | ehealth literacy | Critical/Translational | Critical/Translational | Critical |
| Q11. Identify health information is fake | ehealth literacy | Critical/Translational | Critical/Translational | Critical |
| Q12. Identify a website that is safe for sharing personal health | ehealth literacy | Critical/Translational | Critical/Translational | Critical |
| Q13. Identify information is relevant to health needs | ehealth literacy | Critical/Translational | Critical/Translational | Critical |
| Q14. Know how to evaluate the credibility of others | ehealth literacy | Critical/Translational | Critical/Translational | Critical |
| Q15. Learn to manage health in a positive way | ehealth literacy | Critical/Translational | Critical/Translational | Translational |
| Q16. Use the Internet as a tool to improve health | ehealth literacy | Critical/Translational | Critical/Translational | Translational |
| Q17. Use the information to make a decision | ehealth literacy | Critical/Translational | Critical/Translational | Translational |
| Q18. Learn about topics had relevant to me | ehealth literacy | Critical/Translational | Critical/Translational | Translational |
| Chi-Square | 912.388 | 765.282 | 674.437 | 488.49 |
| Degrees of Freedom | 135 | 134 | 132 | 117 |
| p-value | <0.01 | <0.01 | <0.01 | <0.01 |
| RMSEA | 0.157 | 0.142 | 0.132 | 0.116 |
| (90% CI) | (0.147; 0.166) | (0.132; 0.151) | (0.122; 0.142) | (0.106; 0.127) |
| TLI | 0.780 | 0.82 | 0.843 | 0.897 |
| CFI | 0.806 | 0.842 | 0.865 | 0.907 |

(*RMSEA = root mean square error of approximation. TLI = Tucker-Lewis index. CFI = comparative fit index*)

The results obtained from the Confirmatory Factor Analysis for the TeHLI were estimated in Figure 2. The standardised coefficients range from 0.87 to 1.1. The highest coefficient was found in 4 items (Q4, Q11, Q13, and Q17), while the lowest was Q8.

**Figure 1.**
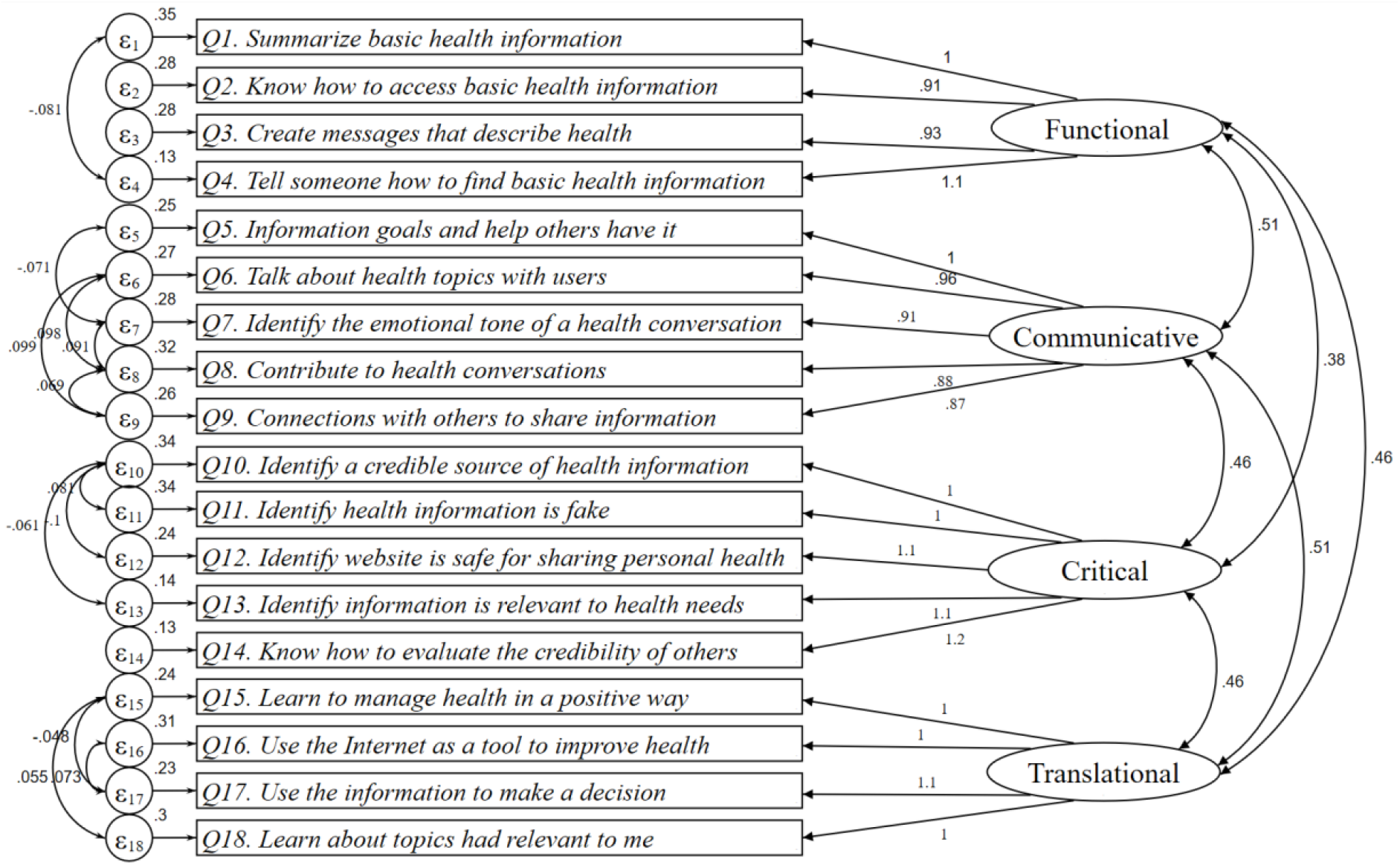
CFA models through structural equation modelling for the Transactional eHealth Literacy Instrument (RMSEA = 0.116; CFI: 0.907; SRMR: 0.047).

The values of ΔCFI were <0.01 when all models, with progressive restrictions, were compared across age groups, education, household economic (Table 4). These groups reported strict invariance. Besides, the configural model of gender, marital status, and location had fit statistics of RMSEA <0.08, CFI>0.9. However, in the metric model, the value of ΔCFI was higher than 0.01 at all three variables (gender, marital status, and location). Metric invariance was therefore not supported by the data.

**Table 4:** Measurement invariance in the Transactional eHealth Literacy questionnaire for the different groups.

| Group | Invariance | x2(df) | x2/df | CFI | RMSEA (90%CI) | $\Delta$ x2($\Delta$ df) | ΔCFI | $\Delta$ RMSEA |
| --- | --- | --- | --- | --- | --- | --- | --- | --- |
| **Gender** | Configural | 288.467 (258) | 1.118 | 0.966 | 0.032 (0.000; 0.050) | - | - | - |
|  | Metric | 289.257 (272) | 1.063 | 0.981 | 0.023 (0.000; 0.044) | 0.790 (14) | 0.015 | 0.009 |
|  | Strong | 307.891 (286) | 1.077 | 0.976 | 0.026 (0.000; 0.045) | 18.634 (14) | 0.005 | 0.003 |
|  | Strict | 326.862 (304) | 1.075 | 0.975 | 0.025 (0.000; 0.045) | 18.971 (18) | 0.001 | 0.001 |
| **Education** | Configural | 406.792 (387) | 1.051 | 0.978 | 0.026 (0.000; 0.049) | - | - | - |
|  | Metric | 436.604 (415) | 1.052 | 0.976 | 0.026 (0.000; 0.048) | 29.812 (28) | 0.002 | 0.000 |
|  | Strong | 466.686 (443) | 1.053 | 0.974 | 0.026 (0.000; 0.048) | 30.082 (28) | 0.002 | 0.000 |
|  | Strict | 501.197 (479) | 1.046 | 0.975 | 0.024 (0.000; 0.046) | 34.511 (36) | 0.001 | 0.002 |
| **Marital status** | Configural | 286.936 (258) | 1.112 | 0.964 | 0.031 (0.000; 0.050) | - | - | - |
|  | Metric | 288.11 (272) | 1.059 | 0.98 | 0.022 (0.000; 0.044) | 1.174 (14) | 0.016 | 0.008 |
|  | Strong | 300.773 (286) | 1.052 | 0.982 | 0.021 (0.000; 0.042) | 12.663 (14) | 0.002 | 0.001 |
|  | Strict | 316.362 (304) | 1.041 | 0.985 | 0.019 (0.000; 0.041) | 15.589 (18) | 0.003 | 0.002 |
| **Age group** | Configural | 278.739 (258) | 1.080 | 0.977 | 0.026 (0.000; 0.046) | - | - | - |
|  | Metric | 282.998 (272) | 1.040 | 0.988 | 0.019 (0.000; 0.042) | 4.259 (14) | 0.010 | 0.008 |
|  | Strong | 297.374 (286) | 1.040 | 0.987 | 0.018 (0.000; 0.041) | 14.376 (14) | 0.001 | 0.001 |
|  | Strict | 314.93 (304) | 1.036 | 0.988 | 0.018 (0.000; 0.040) | 17.556 (18) | 0.001 | 0.000 |
| **Location** | Configural | 415.767 (387) | 1.074 | 0.967 | 0.031 (0.000; 0.052) | - | - | - |
|  | Metric | 419.695 (415) | 1.011 | 0.995 | 0.012 (0.000; 0.042) | 3.928 (28) | 0.028 | 0.019 |
|  | Strong | 450.521 (443) | 1.017 | 0.991 | 0.015 (0.000; 0.042) | 30.826 (28) | 0.003 | 0.003 |
|  | Strict | 484.828 (479) | 1.012 | 0.993 | 0.013 (0.000; 0.041) | 34.307 (36) | 0.002 | 0.002 |
| **Household economy** | Configural | 285.653 (258) | 1.107 | 0.969 | 0.030 (0.000; 0.049) | - | - | - |
|  | Metric | 303.942 (272) | 1.117 | 0.964 | 0.032 (0.000; 0.050) | 18.289 (14) | 0.005 | 0.002 |
|  | Strong | 317.675 (286) | 1.111 | 0.964 | 0.031 (0.000; 0.049) | 13.733 (14) | 0.000 | 0.001 |
|  | Strict | 334.803 (304) | 1.101 | 0.965 | 0.029 (0.000; 0.047) | 17.128 (18) | 0.001 | 0.002 |

## Discussion

This article aimed to validate the TeHLI to see which components of the tool (how many and which components included) would be the best fit statistically and whether the tool applies to groups of different characteristics. Overall, we found that a TeHLI divided into four factors (functional, communicative, critical, and translational) had the best statistical fit (RMSEA = 0.116; CFI = 0.907) and the highest level of internal consistency (Cronbach’s *α* = 0.96). The individual items in the TeHLI have medium-to-high levels of correlation with other items in respective factors (r > 0.6). Upon further analysis, we found partial invariance of the TeHLI - only lacking support for metric invariance.

The study findings substantiate that the most optimal composition of the TeHLI consists of four factors: functional, communicative, critical, and translational eHealth literacy. In this model, Q1 to Q4 measure functional eHealth literacy, Q5 to Q9 measure communicative eHealth literacy, Q10 to Q14 measure critical eHealth literacy, and Q15 to Q18 measure transactional eHealth literacy. This tool composition is in line with the original dimensions of the TMeHL (Paige et al., 2018). The initial measurements of high internal consistency of the four domains by Paige and colleagues (Cronbach’s *α* = 0.91, 0.92, 0.88, and 0.92 respectively) was also supported by our findings (Cronbach’s *α* = 0.88, 0.92, 0.92, and 0.90 respectively) (Paige et al., 2019).

In terms of measurement invariance, the support for configural invariance indicates that the same construct is measured in different population groups. In the context of this study, functional, communicative, critical, and translational eHealth literacy are consistently measured when the TeHLI is deployed in different population groups. The support for scalar invariance indicates the ability to justify the comparison of means across population groups. It indicates that gender, education, marital status, age, location, and household economy do not influence the way participants to respond to the TeHLI to the point that would introduce measurement bias. The support for strict invariance indicates that the residual errors are equivalent across participants. In other words, using the TeHLI across population groups should not produce error margins that substantially differ from each other. However, metric invariance was only found in two groups (education [ΔCFI = 0.002] and household economy [ΔCFI = 0.005]; age group being precisely on the threshold [ΔCFI = 0.010]) and, therefore, only partial metric invariance is supported by this data (Pirralha, 2020), which suggests that the constructs measured have different meanings across some of the participant groups.

Finding partial support for metric invariance is reported to have minimal effects on the mean differences of a latent factor (Steinmetz, 2013). Since two factor loadings reported metric invariance, there are grounds to assume the TeHLI can be used for comparisons across groups (Byrne et al., 1989; Steenkamp & Baumgartner, 1998). Simultaneously, there is also the question of whether partial invariance is sufficient as significant bias can still be introduced into study findings if the partial invariance is ignored (Guenole & Brown, 2014; Steinmetz, 2018). As such, deploying a partially (including invariant items only) and a fully invariant model (including invariant and noninvariant items) and compare the results of interest arises as a viable solution to use the TeHLI across different population groups (Chen, 2008). The risk of this solution is that - if the results differ substantially - there is no clear path forward.

Some limitations apply to this study. The survey findings had a mild-to-moderate negative skewness. However, as extrapolation to the population is impossible given the lack of participants, this limitation does not significantly affect the study findings. The survey was also distributed only among the Vietnamese population. Therefore, the influence of culture could not be determined and needs to be examined in a future study that includes multiple countries or regions.

## Conclusion

Ultimately, the TeHLI can be considered a valuable tool to measure different competency levels of eHealth literacy. As such, the TeHLI has the potential (if deployed correctly) to generate high-quality data that can be used to inform governments and change agents whether the general or a target population is capable of understanding, appraising, distinguishing, and acting appropriately on high- and low-quality health information on the Internet. As a result, it can make vital contributions in informing and guiding policy and practice sustainably into the Digital Age.

## Data Availability

Not available

## Declarations

### Ethical Approval

The protocol of this study was approved by the institutional review board of Youth Research Institute, Vietnam.

### Guarantor

Not applicable

### Contributorship

Giang Thu Vu: conceptualisation, data collection, project management, writing - review & editing,

Thuc Minh Thi Vu: conceptualisation, data collection, project management,

Robin van Kessel: formal analysis, validation, writing - original draft, writing - review & editing,

Brian Li Han Wong: formal analysis, writing - original draft, writing - review & editing,

Tham Thi Nguyen: data collection, data analysis

Thao Phuong Thi Nguyen: data collection, publishing management

Son Hoang Nguyen: data collection

Bach Xuan Tran: conceptualisation, data analysis: review

Carl A. Latkin: writing - review & editing

Cyrus Ho: writing - review & editing

Roger Ho: writing - review & editing

## Acknowledgements

None.

